# Artificial intelligence of arterial Doppler waveforms to predict major adverse outcomes among patients evaluated for peripheral artery disease

**DOI:** 10.1101/2023.07.21.23293024

**Authors:** Robert D McBane, Dennis H. Murphree, David Liedl, Francisco Lopez-Jimenez, Itzhak Zachi Attia, Adelaide M. Arruda-Olson, Christopher G. Scott, Naresh Prodduturi, Steve E. Nowakowski, Thom W. Rooke, Ana I. Casanegra, Waldemar E. Wysokinski, Damon E. Houghton, Haraldur Bjarnason, Paul W. Wennberg

## Abstract

**Background:** Patients with peripheral arterial disease (PAD) are at increased risk for major adverse cardiac (MACE), limb (MALE) events and all-cause mortality. Developing tools capable of identifying those patients with PAD at greatest risk for major adverse events is the first step for outcome prevention. This study aimed to determine whether computer assisted analysis of a resting Doppler waveform using deep neural networks can accurately identify PAD patients at greatest risk for adverse outcome events.

**Methods:** Consecutive patients (4/1/2015-12/31/2020) undergoing ankle brachial index (ABI) testing were included. Patients were randomly allocated to training, validation and testing subsets (60%/20%/20%). Deep neural networks were trained on resting posterior tibial arterial Doppler waveforms to predict MACE, MALE and all-cause mortality at 5 years. Patients were then analyzed in quartiles based on the distribution of each prediction score.

**Results:** Among 11,384 total patients, 10,437 patients met study inclusion criteria (mean age 65.8±14.8 years; 40.6% female). The test subset included 2,084 patients. During 5 years of follow up, there were 447 deaths, 585 MACE and 161 MALE events. After adjusting for age, sex, and Charlson index, deep neural network analysis of the posterior tibial artery waveform provided independent prediction of death (Hazard ratio 2.45 95% confidence interval 1.79-3.36), MACE (HR 1.98, 95%CI 1.50-2.62) and MALE (HR 11.65 95%CI 5.65-24.04) at 5 years with similar results at 1 year.

**Conclusion:** An artificial intelligence enabled analysis of a resting Doppler arterial waveform enables identification of major adverse outcomes including all-cause mortality, MACE and MALE among PAD patients.

## INTRODUCTION

Atherosclerotic peripheral arterial occlusive disease (PAD) affects more than 8 million Americans (1). The clinical presentation of PAD ranges from asymptomatic disease to limb threatened ischemia (2). Patients with PAD are at increased risk for both major adverse cardiac events (MACE) including mortality and major adverse limb events (MALE) including limb amputation. Indeed, PAD is a well-recognized independent predictor for adverse outcomes beyond traditional atherosclerotic risk factors. Despite this, PAD patients are infrequently provided guidelines endorsed risk factor modification (3–9). Less than 20% of patients with PAD have all for major atherosclerotic risk factors addressed (3,4). Compared to coronary artery disease patients, PAD patients are often half as likely to receive statin therapy, antiplatelet agents, tobacco cessation management, or hypertension control to goal (3–6). And yet, high-intensity statin and antiplatelet therapy are known to improve both amputation and 5-year survival rates among patients with limb ischemia (10–12).

Early disease identification with prompt initiation of guideline directed risk management is central to improving outcomes among patients with PAD (8). PAD is often poorly recognized despite patient complaints of exertional leg discomfort and patients with asymptomatic disease are only rarely identified (9,15–17). Delayed diagnosis results in missed opportunities to impact the natural history of disease. In recent years, there has been a near doubling of young patients with PAD, age 18-64, where their first interaction with health care includes hospital admission with chronic limb threatening ischemia (14). These patients had unattended modifiable risk factors including diabetes, nicotine addiction, dyslipidemia and hypertension. By the time patients have presented with chronic limb threatening ischemia, the risk of limb loss, adverse cardiac events and death is substantial (18).

Public health strategies for improving outcomes among patients with PAD must include a system of early disease detection which is affordable, accurate, reproducible, noninvasive, and technically easy to perform. Ideally, data from these testing platforms would go beyond disease detection and provide an assessment of risk for hard outcomes. Noninvasive screening tests for peripheral arterial occlusive disease include the ankle-brachial index (ABI) and Doppler arterial waveform assessment. We have recently shown that artificial intelligence (AI) using deep neural networks can accurately identify patients with PAD based solely on Doppler waveform analysis of the posterior tibial artery (19). Using the same approach, we sought to determine whether an AI algorithm assessment of posterior tibial arterial Doppler signal can accurately identify PAD patients at greatest risk for major adverse cardiac and limb events and all-cause mortality.

## METHODS

### Study Design and patients

Consecutive patients over 18 years of age undergoing clinically indicated lower extremity arterial testing at the Mayo Clinic Gonda Vascular Laboratory between April 8, 2015 – December 31, 2020 were analyzed.

Subjects were excluded who lacked MN research authorization (**Figure 1**). The study was approved by the Mayo Clinic Institutional Review Board.

**Figure 1.**
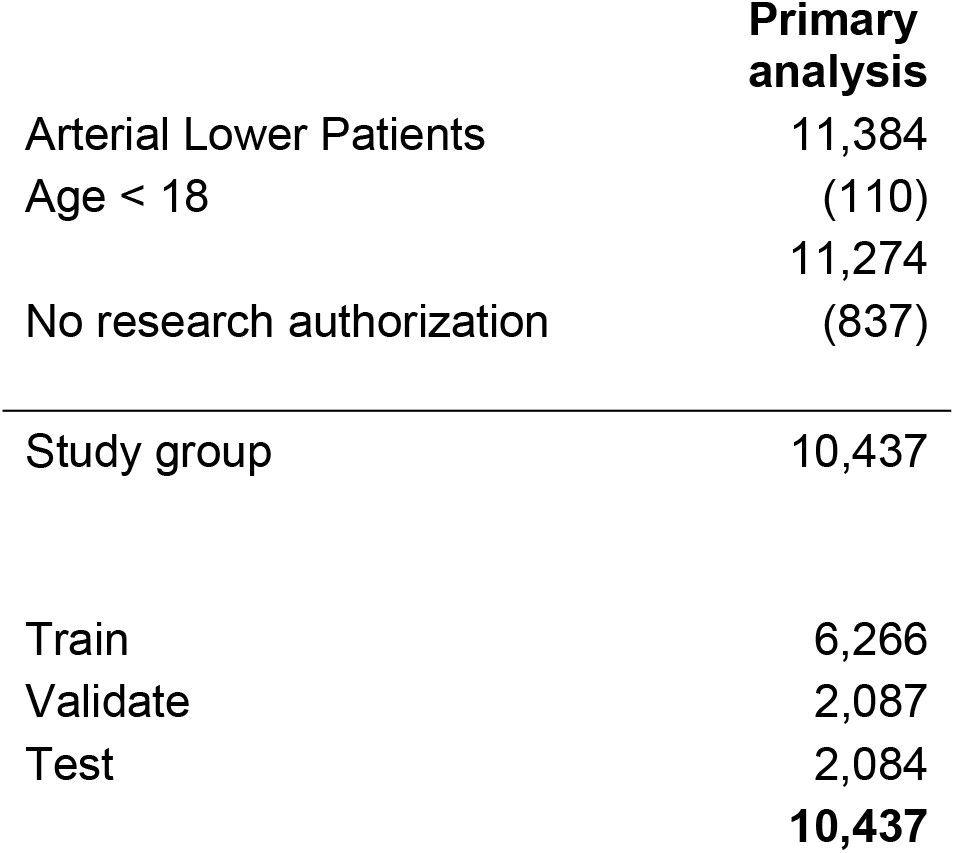
Patient selection and exclusion flow diagram. In the primary study group (2015 – 2020), 11,384 patients had lower extremity arterial study performed. Of these, 110 were excluded due to age cutoff, 837 lacked MN research authorization. The final study group consisted of 10,437 patients which were then divided into train (n=6,266), validation (n=2,087) and test (n=2,084) subgroups.

### Lower extremity arterial test protocol

Arterial disease severity was assessed as previously described (19). Trained technicians dedicated to the Gonda Mayo Clinic Noninvasive Vascular Laboratory performed each evaluation.

With the patient lying supine, continuous wave (CW) Doppler waveforms were recorded at the common femoral, superficial femoral, popliteal, posterior tibial, and dorsalis pedis arterial segments for both legs (Parks Vascular Flo-lab System, Sonova E 2100 SX). Once the CW Doppler signal was identified, waveform sizes were adjusted starting at a gain of 20% until the waveform fit within the monitor display and then archived electronically. Ankle brachial index measurement was performed as previously described (19).

### Outcome Measure

The primary study outcome was the ability of the artificial intelligence (AI) enhanced analysis of resting posterior tibial arterial Doppler waveform to identify all-cause mortality. Secondary outcomes included major adverse cardiac and limb events. Clinical data were collected from a centralized digital data warehouse that contains complete records of all patients evaluated in all sites of the Mayo Clinic enterprise. The Mayo Clinic electronic medical record (EMR) contains details for every inpatient hospitalization, outpatient visit, radiology examination, laboratory and pathology result (including autopsy reports).

A validated algorithmic medical record analysis for identifying patient comorbidities and adverse outcomes was employed using computational phenotyping algorithms which included procedural codes and ICD-9 and ICD-10 diagnosis codes found in the EMR prior to their index date (20–24). Adverse outcomes occurred after the index date. Major adverse cardiac events composite (MACE) included acute myocardial infarction, stroke, coronary revascularization, and all-cause mortality (25).

Major adverse limb event composite (MALE) included acute or chronic limb ischemia, limb revascularization, and major amputations.

### Artificial Intelligence Analysis

The primary dataset consisted of consecutive patients undergoing vascular laboratory testing. For patients with repeat vascular testing, the first study was used. The primary dataset was divided into three subsets, train (n=6266; 60%), validation (n=2087; 20%), and test (n=2084; 20%) via outcome stratified random sampling. Each patient was uniquely assigned to a single group. The validation set was used to monitor performance during training and for alternative classification threshold determination, while the test set was used for independent evaluation.

Separate models were trained for each of the hard outcomes including all-cause mortality, MACE, MALE, and combined MACE plus MALE. These outcomes were assessed separately using events that occurred within one and five years of the index study date. For patients with multiple events, only the first event was considered. For analyses of MALE and combined MACE and MALE, patients with prior limb events were excluded. In order to take time into account, these analyses used weights based on the redistribute to the right algorithm (RTTR)(26). Specifically, each subject was given a weight for each outcome and the model development dataset replicated patients proportionate to the RTTR weight for that particular analysis.

A variety of deep neural network (DNN) architectures were explored; all trained as binary classifiers on the resting Doppler waveforms to predict hard outcomes.

Feature scaling of the input Doppler data was accomplished by dividing by 128. All models used were 1-dimensional convoluted neural networks (CNNs) with an InceptionTime architecture (19). Doppler data from left and right posterior tibial (**PT**) arteries were fed into the InceptionTime network, and outputs prior to the final flobal average pooling layer were merged via simple addition then passed to a fully connected output layer. The activation function of all neurons was the rectified linear unit (ReLU). The softmax function was used as the activation function of the output layer. Training was performed by minimizing the binary cross-entropy loss, with weights from both the InceptionTime and final output layers updated during each pass. A batch size of 64 samples was used and models were trained for 100 epochs. Best model weights from checkpoints were chosen based on validation loss. Class weights were introduced to address the imbalance between classes (weights inversely proportional to class frequency). Additional hyperparameters such as dropout, and learning rate were also optimized using a grid search.

### Statistical Analysis

Clinical measurements and descriptive characteristics were summarized using frequencies for categorical variables and mean and standard deviation, median and quartiles for continuous variables.

After development of predictions for each outcome, patients were grouped into quartiles based on the distribution of the prediction score in the training dataset. Thus, patient group assignment could vary by outcome of interest. Kaplan-Meier analysis was used to plot survival free of each outcome by corresponding prediction quartile. Cox proportional hazards regression was used to estimate the hazard ratios (HR) and associated 95% confidence intervals (CI) with and without adjustment for age, sex, and Charlson comorbidity index. A measure of discrimination ability of each model, the survival concordance index (C-index) and 95% CI, was also estimated with and without adjustments. These analyses were done separately for each dataset and each event type at 1 and 5 year time intervals.

In all cases, a two-tailed p-value of less than 0.05 was considered statistically significant. Statistical analysis was done using SAS statistical software (SAS version 9.4; SAS Institute Inc.) and the R software package v3.6.2 (Team R. Core R: A Language and Environment for Statistical Computing, Version 3.5. 3. Vienna: R Foundation for Statistical Computing 2019). Neural network development was performed using Tensorflow 2.2.0 and Python 3.6.12.

## RESULTS

### Demographic Characteristics

Of the original cohort of 11,384 patients being evaluated in this laboratory over the study dates, 110 were excluded due to age < 18 years (**Figure 1**). An additional 837 subjects were excluded due to lack of research authorization.

The final cohort included 10,437 patients with a mean age of 65.8 years (SD 14.8 years); 40.6% were female (**Table 1**). Cardiovascular risk factors were prevalent amongst this cohort including hypertension (49.5%), dyslipidemia (56.4%), diabetes mellitus (40.0%), and any history of tabacco use(63.2%). Coronary artery disease, prior myocardial infarction, prior coronary artery bypass grafting, and heart failure were less frequent involving 20% or fewer patients. Atrial fibrillation, prior stroke, emphysema, and chronic kidney disease were additional though less frequent clinical variables of interest.

**Table 1.** Patient Demographics.

| **Table 1. Patient Demographics** |  |  |  |  |
| --- | --- | --- | --- | --- |
| **Variable** | **Train**<br>**(n=6,266)** | **Validation**<br>**(n=2,087)** | **Test**<br>**(n=2,084)** | **Total**<br>**(n=10,437)** |
| **Age years, Mn (SD)** | 65.9 (14.7) | 65.2 (15.2) | 66.1 (15.0) | 65.8 (14.8) |
| **Female Gender, n (%)** | 2515 (40.1%) | 877 (42.0%) | 836 (40.1%) | 4228 (40.5%) |
| **Hypertension, n (%)** | 3157 (50.4%) | 1028 (49.3%) | 1032 (49.5%) | 5217 (50.0%) |
| **Dyslipidemia, n (%)** | 3557 (56.8%) | 1174 (56.3%) | 1170 (56.1%) | 5901 (56.5%) |
| **Diabetes Mellitus, n (%)** | 2524 (40.3%) | 831 (39.8%) | 856 (41.1%) | 4211 (40.3%) |
| **Tobacco History, n (%)**<br>**(n=10,086)** | 3847 (63.7%) | 1277 (63.2%) | 1280 (63.2%) | 6404 (63.5%) |
| **Coronary Artery Disease, n (%)** | 1295 (20.7%) | 440 (21.1%) | 442 (21.2%) | 2117 (20.9%) |
| **Prior Myocardial Infarction, n (%)** | 877 (14.0%) | 301 (14.4%) | 288 (13.8%) | 1466 (14.0%) |
| **Prior Coronary Bypass Surgery, n (%)** | 1058 (16.9%) | 355 (17.0%) | 340 (16.3%) | 1753 (16.8%) |
| **Heart Failure, n (%)** | 1237 (19.7%) | 427 (20.5%) | 387 (18.6%) | 2051 (19.7%) |
| **Atrial Fibrillation, n (%)** | 1193 (19.0%) | 400 (19.2%) | 397 (19.0%) | 1990 (19.1%) |
| **Prior Stroke, n (%)** | 801 (12.8%) | 262 (12.6%) | 285 (13.7%) | 1348 (12.9%) |
| **Emphysema, n (%)** | 1065 (17.0%) | 365 (17.5%) | 356 (17.1%) | 1786 (17.1%) |
| **Chronic Kidney Disease, n (%)** | 1238 (19.8%) | 389 (18.6%) | 383 (18.4%) | 2010 (19.3%) |
| **Charlson Index, median (IQR)** | 3.0 (1.0, 6.0) | 3.0(1.0, 6.0) | 3.0 (1.0, 6.0) | 3.0 (1.0, 6.0) |

### Model Development

Model development was performed on the training set (N=6266) and the validation set (N=2087) was used for model tuning. For each outcome, the quartiles of the distribution of prediction scoes were extracted from the training set. These quartile cutpoints were used to divide patients into groups in the training set as well as the validation and test sets (N=2084). Each of the outcomes were plotted by quartile using the Kaplan-Meier method. Within the Train and Validation datasets, event rates varied based on prediction quartile. When outcomes were evaluated in the Train dataset of 6,266 patients, the 5 year death rate in the first quartile was 14% (11-16) compared to 60% (56-64) in the fourth quartile (**Figure 2A**). The 1 and 5 year rates (95% Confidence Intervals) of MACE, MALE, and MACE+MALE were 28% (24-32), 0% (0-0), and 35% (30-40) in the first quartile compared to 77% (73-81), 58% (48-66), and 82% (77-85) in the fourth quartile, respectively (**Table 2**). When evaluated in the validation set, similar differences in rates, HR, and C-Indices across quartiles were noted.

**Figure 2.**
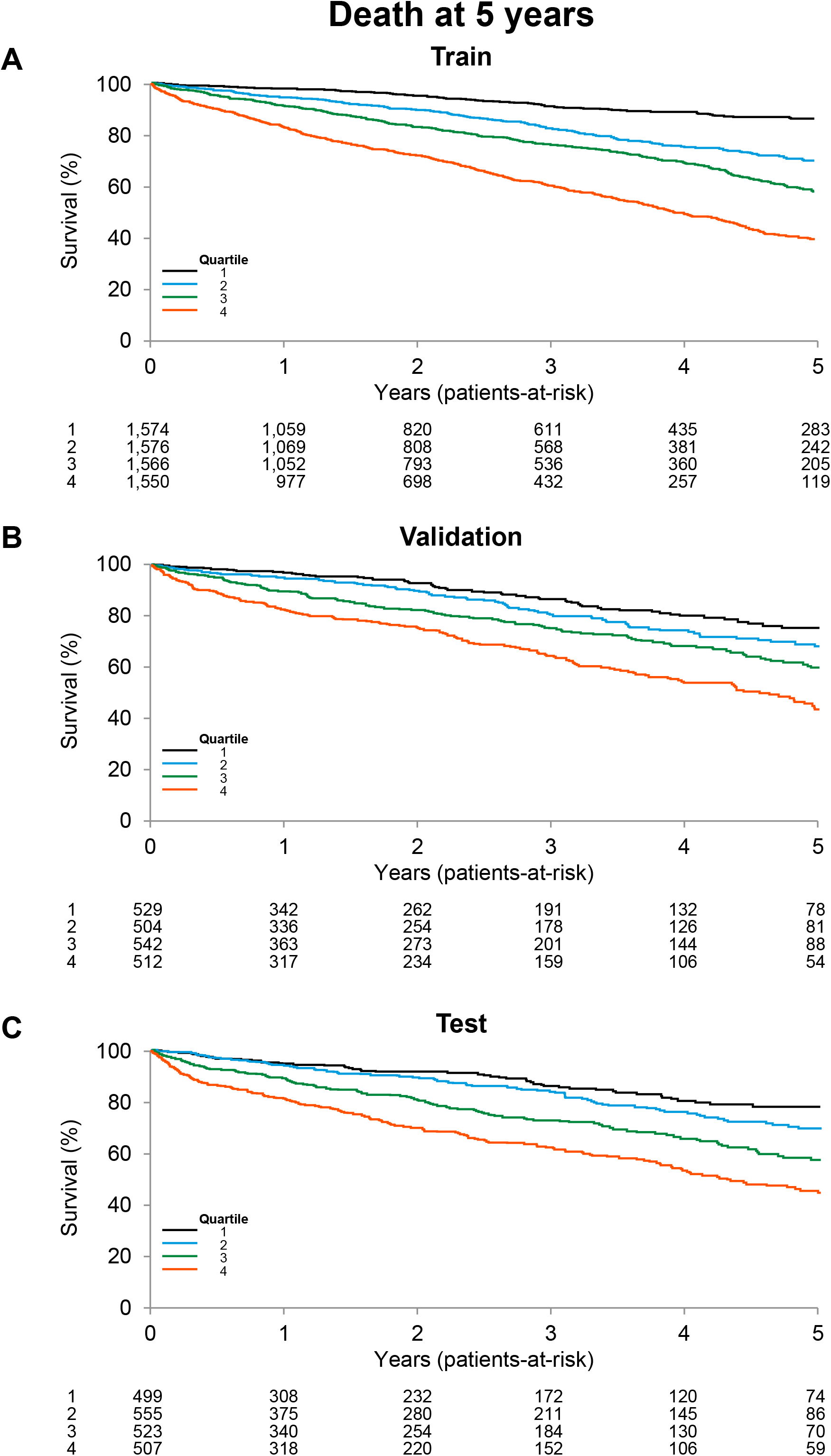
Model train, validation, and test performance for all-cause mortality at 5 years. Patients were divided into quartiles based on the distribution of the prediction score taken from the training set. Over the 5-year follow up, there were 2,033 deaths. Survival plots are provided for each quartile for the train (**panel A**), validation (**panel B**), and test subsets (**panel C**).

**Table 2.** Tain and Validation Dataset 5 year Outcomes.

| Dataset,<br>1/5 year KM<br>event rate (%) | Quartile based on 5 year prediction |  |  |  | Hazard Ratio<br>(95% CI)<br>Q4 vs. Q1 | C-Index<br>(95% CI) |
| --- | --- | --- | --- | --- | --- | --- |
|  | 1 | 2 | 3 | 4 |  |  |
| Train |  |  |  |  |  |  |
| Overall Mortality | 2<br>14 | 5<br>30 | 9<br>42 | 17<br>60 | 5.88<br>(4.84-7.14) | 0.67<br>(0.65-0.69) |
| Major Adverse Cardiac Events (MACE) | 10<br>28 | 13<br>46 | 21<br>63 | 31<br>77 | 3.90<br>(3.33-4.55) | 0.63<br>(0.62-0.65) |
| Major Adverse Limb Events (MALE) | 0<br>0 | 0<br>0 | 5<br>11 | 21<br>58 | 226<br>(56-909) | 0.83<br>(0.80-0.86) |
| MACE plus MALE | 11<br>35 | 13<br>47 | 22<br>60 | 37<br>82 | 3.72<br>(3.19-4.35) | 0.65<br>(0.63-0.66) |
| Validation |  |  |  |  |  |  |
| Overall Mortality | 3<br>25 | 6<br>32 | 10<br>40 | 18<br>57 | 3.19<br>(2.40-4.23) | 0.63<br>(0.60-0.66) |
| Major Adverse Cardiac Events (MACE) | 10<br>32 | 15<br>54 | 21<br>55 | 27<br>71 | 2.82<br>(2.17-3.66) | 0.60<br>(0.57-0.62) |
| Major Adverse Limb Events (MALE) | 1<br>4 | 2<br>9 | 7<br>21 | 21<br>42 | 15.9<br>(7.4-34.3) | 0.76<br>(0.71-0.81) |
| MACE plus MALE | 11<br>39 | 15<br>45 | 24<br>59 | 37<br>79 | 3.22<br>(2.47-4.19) | 0.64<br>(0.61-0.66) |

### All-Cause Mortality

For the primary outcome, 5 year all-cause mortality was evaluated in the test dataset comprised of 2,084 patients. When patients were divided into quartiles determined from the distribution of the prediction score taken from the training set, results were similar to the train and validation sets (**Table 3**). Within the test dataset 447 patients died within 5 years and the 5 year mortality rates varied from 22% (16-27) in the first quartile to 56% (49-61) in the fourth quartile (**Table 3**, **Figure 2C).** Similar to the training and validation datasets, cardiovascular risk factor prevalence and co-morbidities were noted and differed by patient quartile (**Table 2S**). After adjusting for age, sex, and Charlson Comorbidity Index, having an AI Doppler signal prediction in the fourth quartile remained a significant predictor of mortality (HR 2.45, 95%CI 1.79 – 3.36) compared to patients in quartile 1. The combined model C-index improved from 0.64 (0.61-0.67) to 0.72 (0.70-0.75) with the inclusion of age, sex, and Charlson Comorbidity Index.

**Table 3.** Test Dataset 5 year Outcomes.

| **Test Dataset,**<br>**1/5 year KM**<br>**event rate (%)** | **Quartiles** |  |  |  | **Hazard Ratio**<br>**(95% CI)**<br>**Q4 vs. Q1** | **C-Index**<br>**(95% CI)** |
| --- | --- | --- | --- | --- | --- | --- |
|  | **1** | **2** | **3** | **4** |  |  |
| Overall<br>Mortality | 5<br>22 | 6<br>31 | 11<br>43 | 19<br>56 | 3.56<br>(2.64-4.81) | 0.64<br>(0.61-0.67) |
| Major Adverse<br>Cardiac Events<br>(MACE) | 10<br>36 | 13<br>48 | 23<br>58 | 31<br>70 | 2.81<br>(2.15-3.68) | 0.62<br>(0.59-0.64) |
| Major Adverse<br>Limb Events<br>(MALE) | 1<br>6 | 3<br>10 | 6<br>18 | 21<br>38 | 11.71<br>(5.91-23.21) | 0.75<br>(0.71-0.80) |
| MACE plus<br>MALE | 10<br>35 | 13<br>48 | 26<br>65 | 39<br>65 | 3.27<br>(2.46-4.35) | 0.65<br>(0.62-0.68) |

### Major Adverse Cardiac Events

During 5 years of follow up, 585 patients experienced a major adverse cardiac event (MACE). Events included 114 myocardial infarctions, 349 strokes, and 122 cardiac bypass graft procedures. Five year event rates differed by patient quartile ranging from 36% (27-45) in the first quartile to 70% (62-75) in the fourth quartile (**Figure 3A**, **Table 3**). After adjusting for age, sex, and Charlson Index, AI Doppler signal analysis remained a significant predictor of MACE (HR 1.98, 95%CI 1. 50 – 2.62; C-Index 0.67, 95% CI 0.65 – 0.70).

**Figure 3.**
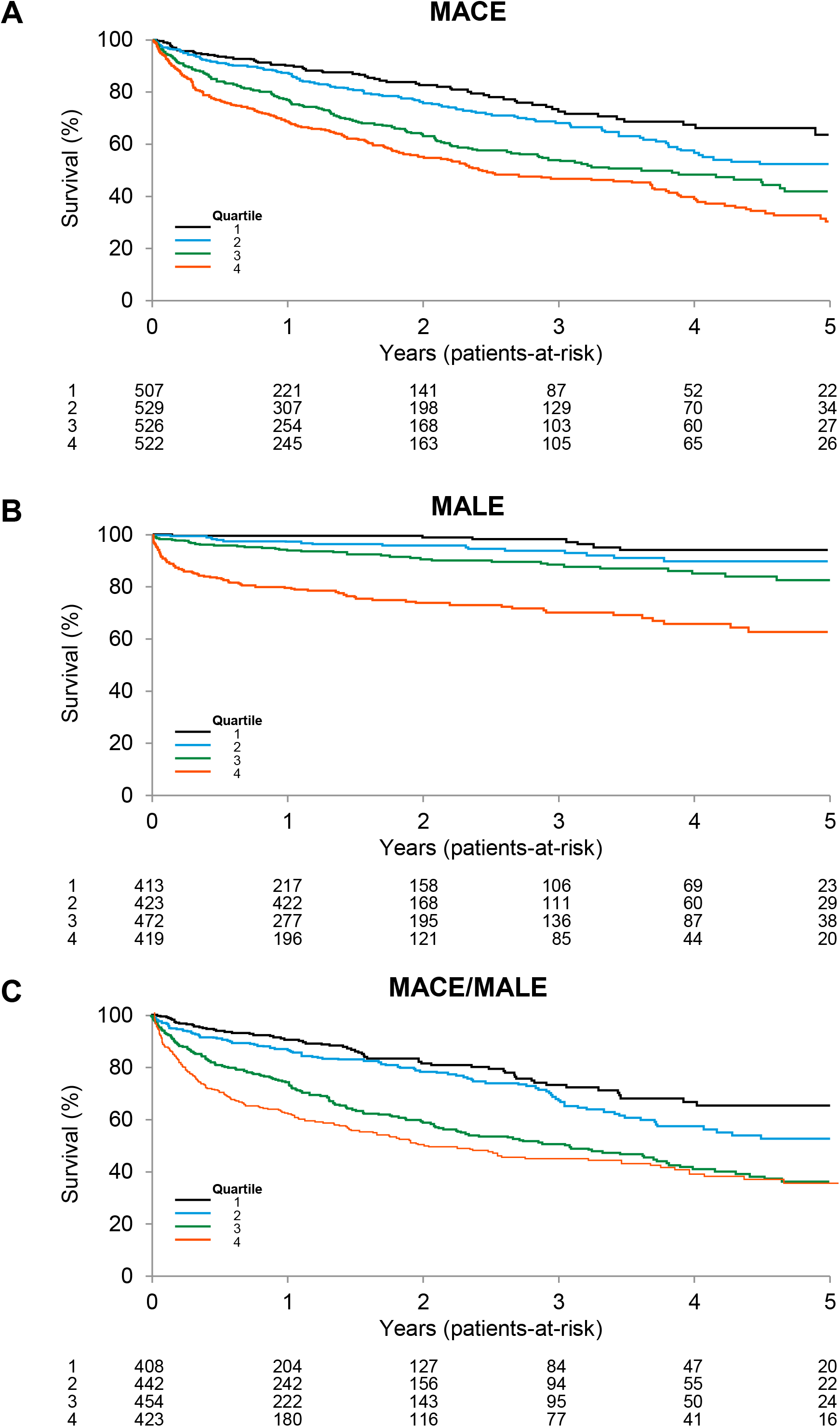
Model performance for major adverse cardiac and limb events at 5 years. The 5 year outcomes are provided for major adverse cardiac events (**MACE, panel A**), major adverse limb events (**MALE, panel B**), and combined major adverse events (**MACE plus MALE, panel C**).

### Major Adverse Limb Events

During 5 years of follow up, 161 patients had a major adverse limb event (MALE) in the subset of 1727 patients without prior major adverse limb events. This included 1 major limb amputation, 158 acute limb ischemic events, and 2 limb revascularization procedures. Five year event rates differed by patient quartile ranging from 6% (2-10) in the first quartile to 38% (29-45) in the fourth quartile (**Figure 3B**, **Table 3**). After adjusting for age, sex, and Charlson Index, AI Doppler signal analysis remained a significant predictor of MALE (HR 11.65, 95%CI 5.65 – 24.04; C-Index 0.78, 95% CI 0.73 – 0.83).

### Combined Major Adverse Cardiac plus Limb Events

These combined events were assessed over 5 years (**Figure 3C**, **Table 3**). After adjusting for age, sex, and Charlson Index, AI Doppler signal analysis remained a significant predictor of combined events (HR 2.64, 95%CI 1.97 – 3.54; C-Index 0.69, 95% CI 0.67 – 0.72).

### Outcomes at 1 year

The hard outcomes including all cause mortality, major adverse cardiac and limb events were analyzed at one year from the index vascular laboratory test date. These data are found in the Supplementary files (**Figure 1S**, **Table 1S**). After adjusting for age, sex, and Charlson index, the AI analysis remained significant for all-cause mortality (HR 1.68 [1.05 – 2.71]; C-index 0.73 [0.69-0.77]), MACE (HR 2.13 [1.50 – 3.03]; C-index 0.68 [0.65-0.71]), MALE (HR 10.09 [4.06 – 25.10]; C-index 0.79 [0.73-0.85]), and combined MACE+MALE (HR 3.41 [2.35-4.93]; C-index 0.72 [0.69-0.75]).

## DISCUSSION

The main findings of the current study include the finding that a deep neural network can help predict hard outcomes including all-cause mortality, major adverse cardiac and limb events based solely on the resting arterial Doppler waveform analysis of the posterior tibial artery. After adjusting for age, sex, and major co-morbidities encompassed in the Charlson index, the machine learning analysis of the Doppler waveform identified patients at risk for all-cause mortality at 5 years with a HR of 2.45. Similarly, the model retrained for other hard outcomes identified those at risk for major adverse cardiac events (HR 1.98) and major adverse limb events (HR 11.65). The adjusted AI analyses were similarly strong for all-cause mortality at one year (HR 1.68), MACE (HR 2.13), and MALE (HR 10.09).

These findings may have relevant patient management implications. First, such findings may incentivize cardiovascular risk factor treatment implementation. Alerting the patient, family members and health care provider regarding hard outcome risk may prompt early implementation and adherence to aggressive risk factor modification.

Arguably, one of the biggest hurdles is compliance with smoking cessation programs. Knowledge of third or fourth quartile risk positioning may promote patient buy-in to participation. Dietary and lipid management, hypertension control and improved diabetes management may all be facilitated with this knowledge. Second, both the implementation and third party payment of aggressive lipid modification with drugs such as PCSK9 inhibitors, bempedoic acid, ezetimibe, or inclisiran added to maximally tolerated statins might be facilitated for those at highest risk (27). Third, knowledge of patient risk for these outcomes may prompt a strategy of more aggressive clinical follow up and coaching in an effort to improve outcomes. For these combined reasons, machine learning analysis of Doppler waveforms may offer a new tool for widespread evaluation and management of patients with PAD.

These findings build on previous work using machine learning to identify patients with PAD and assess outcomes (28). A deep neural network analysis of posterior tibial artery Doppler waveforms was used to identify PAD among a cohort of 3,432 patients undergoing rest and post-exercise ankle brachial index testing at Mayo Clinic Gonda Vascular Laboratory (19). Artificial intelligence enabled analysis of resting Doppler arterial waveforms identified patients with peripheral artery disease with an area under the receiver operating characteristic curve (AUC) of 0.94, sensitivity 0.83, specificity 0.88, accuracy 0.85, positive predictive value (PPV) 0.90 and negative predictive value (NPV) 0.80. In a second study using a validated electronic algorithm for PAD detection, a community-based inception cohort of 1676 Olmsted County residents with PAD was identified (23). A prognostic model of clinical variables was used to stratify patients into risk categories for 5-year mortality outcomes. For the highest risk quartile, the hazard ratio was 8.44 (95%CI 6.66-10.70) compared to the lowest risk quartile with a hazard ratio of 0.35 (95%CI 0.21-0.58). In a third study, demographic, clinical and genomic factors were used to construct a machine learning algorithm to identify PAD and predict risk of mortality (29).

Among a cohort of 1047 participants of the Gene PAD study, a random forest model predicted mortality with an area under the curve of 0.76 (95% CI 0.68 – 0.84). Others have used machine learning to predict outcomes among patients with other cardiovascular phenotypes. Weichwald et al. developed a novel scoring system for assessing one year outcomes following acute coronary syndromes among a cohort of 2,168 patients (30). This tool, coined the SPUM-ACS score, compiled 8 clinical and laboratory variables including a history of peripheral artery disease, predicted 1 year all-cause mortality with area under the curve of 0.86 (95%CI 0.83-0.89) which outperformed the GRACE 2.0 score. Similar tools have been developed for carotid disease and ischemic stroke (31,32).

Beyond mortality and major adverse cardiac events, machine learning has been used to determine major adverse limb events following revascularization procedures. Among 327 patients with PAD undergoing endovascular therapy, a deep neural network algorithm including clinical, demographic, and imaging variables predicted major adverse limb events with an area under the receiver operator curve of 0.80 (95%CI 0.68 – 0.89) which significantly outperformed logistic regression analysis models (33). These findings add to other risk assessment tools for early and 1-year outcomes following revascularization procedures (34–37).

The artificial intelligence tool described in this manuscript has several unique advantages whereby both the 1 and 5 year risk for major adverse limb events can be estimated well before chronic limb threatening ischemia occurs based solely on deep neural assessment of a posterior tibial arterial waveform Doppler signal. The 1-year time projection provides an opportunity for timely vascular subspecialty interactions, while ideally preventing major limb loss. The 5-year horizon provides sufficient time for risk modification to potentially alter disease trajectory. With automated harvesting of clinical variables from the electronic health record, a future state might be envisioned where best practice clinical decision support systems can be triggered real-time upon Doppler waveform acquisition to a prediction model. Combined with guideline risk modification recommendations to inform clinical decision making, prescribing patterns might be facilitated to improve outcomes for these patients.

This study is best understood in the context of its limitations. First, the analysis requires a careful recording of the posterior tibial artery Doppler signal. Improper collection of this tracing by untrained personnel may limit the utility of this methodology. Future studies to assess the tolerance of such signal acquisition would be important.

Second, limited racial diversity of our patient population may limit generalizability. Third, the retrospective nature of the study may also limit generalizability. Fourth, referral bias likely influenced subject inclusion. Only patients referred to the Gonda Vascular Center for ankle brachial index assessment were enrolled. This restricts generalizability to the general population.

In conclusion, applying artificial intelligence via deep neural networks to an easy to perform, non-invasive ultrasound measure, may enable the Doppler signal to serve as a potential risk assessment tool for future adverse outcomes including all-cause mortality, major adverse cardiac and limb events. Further validation studies are required to assess test accuracy and reproducibility in community settings outside of a large volume academic vascular laboratory.

## Data Availability

The data will not be made available for public use at this time.

## Notes

### Competing Interest Statement

The authors have declared no competing interest.

### Funding Statement

No external funding.

### Author Declarations

Mayo Clinic IRB

